# Development and clinical application of a rapid and sensitive loop-mediated isothermal amplification test for SARS-CoV-2 infection

**DOI:** 10.1101/2020.05.20.20108530

**Authors:** Xuejiao Hu, Qianyun Deng, Junmin Li, Jierong Chen, Zixia Wang, Xiqin Zhang, Zhixin Fang, Haijian Li, Yunhu Zhao, Pan Yu, Wenmin Li, Xiaoming Wang, Shan Li, Lei Zhang, Tieying Hou

## Abstract

**Background:** The SARS-CoV-2 outbreak urgently requires sensitive and convenient COVID-19 diagnostics for the containment and timely treatment of patients. We aimed to develop and validate a novel reverse transcription loop-mediated isothermal amplification (RT-LAMP) assay to detect SARS-CoV-2 in qualified laboratories and point-of-care settings.

**Methods:** Patients with suspected COVID-19 and close contacts were recruited from two hospitals between Jan 26 and April 8, 2020. Respiratory samples were collected and tested using the RT-LAMP assays, and the results were compared with those obtained by RT-qPCR. Samples yielding inconsistent results between these two methods were subjected to next-generation sequencing for confirmation. The RT-LAMP assay was also tested on an asymptomatic COVID-19 carrier and patients with other respiratory viral infections.

**Results:** Samples were collected from a cohort of 129 cases (329 nasopharyngeal swabs) and an independent cohort of 76 patients (152 nasopharyngeal swabs and sputum samples). The RT-LAMP assay was validated to be accurate (overall sensitivity and specificity: 88.89% and 99.00%; positive and negative predictive values: 94.74% and 97.78%, respectively) and diagnostically useful (positive and negative likelihood ratios: 88.89 and 0.11, respectively). RT-LAMP showed an increased sensitivity (88.89% vs 81.48%) and high consistency (kappa 0.92) compared with RT-qPCR for SARS-CoV-2 screening while requiring only constant temperature heating and visual inspection. The time required for RT-LAMP was less than 1 h from sample preparation to result. In addition, RT-LAMP was feasible for use with asymptomatic patients and did not cross-react with other respiratory pathogens.

**Conclusion:** The developed RT-LAMP assay offers rapid, sensitive and straightforward detection of SARS-CoV-2 infection and could aid the expansion of COVID-19 testing in the public domain and hospitals.

## INTRODUCTION

The skyrocketing COVID-19 outbreak has become a public health emergency of international concern. A total of 4,618,821 confirmed cases and 311,847 deaths have been reported in 216 countries as of May 18, 2020, since early December of 2019, according to the WHO COVID-19 report.^1^ At present, there are still no effective drugs or vaccines reported for COVID-19, and prompt diagnosis, close contact tracking and quarantine management are the hallmarks for the containment of this new pandemic.

Early and accurate diagnosis of SARS-CoV-2 infection is crucial to prevent virus transmission and provide appropriate treatment for patients. Due to its nonspecific symptoms and radiological features overlapping with those of the common cold and influenza, the confirmation of SARS-CoV-2 infection entirely depends entirely on viral RNA detection. ^2–3^ RT-qPCR is the standard and most widely used method for SARS-CoV-2 RNA detection in clinical laboratories. ^4^ Despite its outstanding analytical performance, RT-qPCR-based approaches to detect COVID-19 still suffers from many limitations, such as long turnaround times (2 to 4 h), poor availability (it is currently restricted to public health laboratories), the need for expensive instrumentation, and a high proportion of false negatives or equivocal values (up to 38%) ^5–6^ in upper respiratory samples due to insufficient viral materials. These limitations make the RT-qPCR test far from adequate to meet the current challenge of a tremendous undocumented infected population, asymptomatic transmission ^7^ and convalescence with viral RNA conversion, ^8^ highlighting the pressing need for a more rapid, simple and sensitive approach to quickly identify infected patients in different settings.

Loop-mediated isothermal amplification (LAMP) is regarded as a promising point-of-care test (POCT) due to its advantages of high sensitivity and specificity, rapid reaction and low laboratory infrastructure requirements. ^9^ Reverse transcription LAMP (RT-LAMP) is a type of LAMP method used to detect target RNA with the AMV reverse transcriptase. This approach allows reverse transcription and DNA amplification to be rapidly accomplished at a 60-65 °C constant temperature in less than an hour and in one step, and detailed amplification mechanisms have been previously reported. ^10^ RT-LAMP results can be detected by visual turbidity or fluorescence in real time, which makes this method a practical near-patient assay. In recent years, RT-LAMP has been widely used in specialized laboratory testing as well as field surveys to identify various pathogens, including *Mycobacterium tuberculosis,* ^11^ Zika virus, ^12^ MERS-CoV, ^13^ and SARS-CoV. ^14^ Shirato et al. ^13^ reported the development of a useful RT-LAMP assay for the diagnosis of MERS that was developed in this way, with a detection limit of 3.4 copies per reaction and no cross-reactivity with other respiratory viruses. In addition, Hong et al. ^14^ developed a real-time quantitative RT-LAMP assay for the early and rapid diagnosis of SARS-CoV that demonstrated 100-fold greater sensitivity than conventional RT-qPCR assays.

To accelerate clinical diagnostic testing for COVID-19, we conducted a prospective cohort study to develop and validate a novel RT-LAMP assay capable of detecting SARS-CoV-2 RNA for potential use in centralized facilities and point-of-care settings. Moreover, we compared RT-qPCR and RT-LAMP using clinical samples and demonstrated that RT-LAMP produced a higher sensitivity and cost effectiveness for SARS-CoV-2 detection. To the best of our knowledge, this study is the first to comprehensively assess a rapid RT-LAMP test for both COVID-19 patients and an asymptomatic carrier, the results of which demonstrated it to have improved diagnostic value over current diagnostics for SARS-CoV-2 infection.

## MATERIALS AND METHODS

This study was designed as a prospective observational cohort study with three sequential phases. In the initial stage, we developed a visual and rapid RT-LAMP assay for SARS-CoV-2 detection and assessed its anti-cross interface ability, stability and detection limit. Subsequently, we evaluated the RT-LAMP and standard RT-qPCR assays on 329 nasopharyngeal swabs from a cohort of 129 suspected COVID-19 patients and on the serial upper respiratory samples from an asymptomatic carrier, and the insistent samples between RT-LAMP and RT-qPCR were further subjected to next-generation sequencing (NGS) for SARS-CoV-2 confirmation. Finally, we analyzed an additional 20 patients with other viral infections, 20 healthy individuals, and an independent cohort of 76 cases suspected of having COVID-19 to further validate the detective captivity of RT-LAMP for SARS-CoV-2. The overall study strategy is shown in Figure 1.

**Figure 1.**
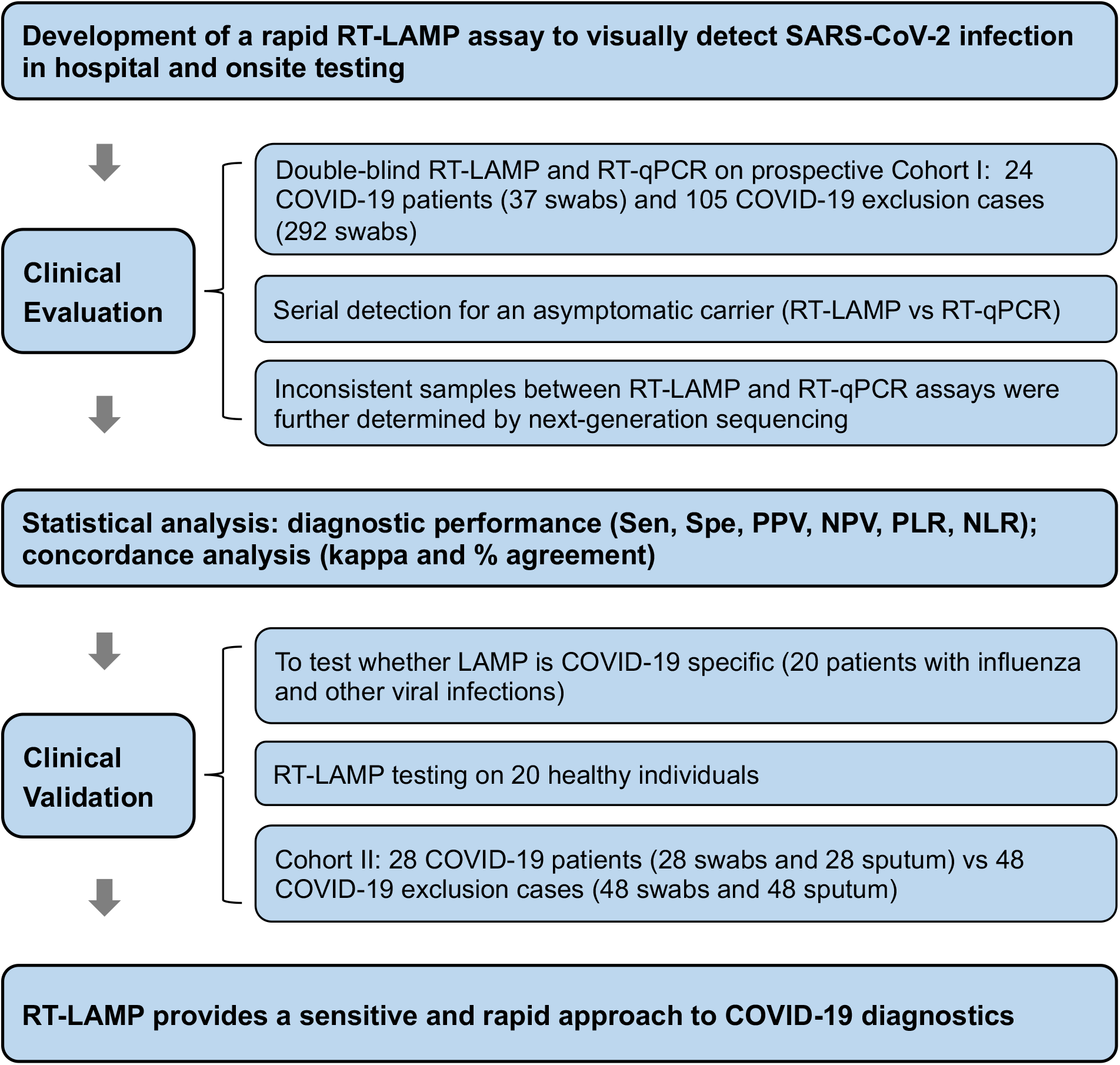
Overview of the study design. Notes: Sen, sensitivity; Spe, specificity; PPV, positive predictive value; NPV, negative predictive value; PLR, positive likelihood ratio; NLR, negative likelihood ratio.

## Subjects and sample enrollment

*Cohort I* Inpatients with clinical-radiological suspicion of COVID-19 presenting to Guangdong Provincial People’s Hospital between January 26 to April 8, 2020 were eligible for inclusion. Close contacts with exposure to confirmed COVID-19 cases were simultaneously enrolled in the present study. Every participant underwent a standard SARS-CoV-2 set of investigations to test COVID-19. The patients’ demographic, clinical, laboratory and radiological findings were collected from their medical records. Serial nasopharyngeal swabs were collected from patients during hospitalization and close contact screening. The sample sizes for swabs were defined by their availability. At least one nasopharyngeal swab from suspected COVID-19 patients was simultaneously sent to the CDC for double-checking as required, where RT-qPCR was standardly utilized for SARS-CoV-2.

The diagnosis of COVID-19 patient was based on acute respiratory infection syndromes and/or the presence of chest imaging features consistent with viral pneumonia accompanied by conformation of positive RT-qPCR test results for SARS-CoV-2 by the CDC, according to the criteria published in the updated COVID-19 Diagnostic Criteria, 7th Edition, China. Suspected COVID-19 patients from Guangdong Provincial People’s Hospital were defined as Cohort I in this study and classified into two groups: COVID-19 and non-COVID-19. COVID-19 patients were further classified as nonsevere cases and severe cases; nonsevere cases included mild and moderate patients with mild pneumonia, and severe cases indicated severe and critically severe patients with acute respiratory distress syndrome (ARDS) or oxygen saturation at rest <93% who required mechanical ventilation or ICU monitoring. ^2^

*Cohort II* We enrolled an independent cohort of suspected COVID-19 patients from Guangdong Second Provincial General Hospital for validation. SARS-CoV-2 testing and the diagnosis procedure for COVID-19 were identical in the two hospitals. A nasopharyngeal swab and 5 mL of morning sputum were collected from suspected COVID-19 patients to validate the diagnostic performance of RT-LAMP for SARS-CoV-2.

In addition, nasopharyngeal swab samples obtained from 20 healthy subjects and 20 patients with other respiratory viral infections were used to test the specificity of RT-LAMP for SARS-CoV-2 detection.

## RNA extraction

Swabs were preserved in 500 μL of virus preservation solution (TianLong, China), which inactivates viruses and preserves all RNA in the specimen. The sputum samples were preprocessed by a standard NALC-NaOH digestion. Total RNA was extracted from specimens within 2 h of collection using a magnetic bead-based viral RNA isolation kit with a DA3200 system instrument (Daan Gene, China) according to the manufacturer’s instructions. The extracts were stored at −70°C until use. RNA extracted from each specimen was tested for SARS-CoV-2 in parallel by RT-qPCR and RT-LAMP in a double-blind manner in a biosafety level 2 laboratory. The samples yielding inconsistent results between these two methods were further analyzed by NGS for verification.

## RT-qPCR amplification

RT-qPCR was performed using an officially approved clinical RT-qPCR kit for the ABI COVID-19 QuantStudio Dx^TM^ real-time PCR system (Applied Biosystems, USA) following the manufacturer’s protocol (Daan Gene). Primer and probe sets targeting the *ORF1ab* and *N* genes of SARS-CoV-2 are provided in Table S1. For RT-qPCR, each 25-μL reaction comprised 17 μL of reaction buffer, 3 μL of enzyme solution, and 5 μL of template RNA. The cycling program started at 50°C for 15 min for reverse transcription, followed by 95°C for 15 min for PCR initial activation and 45 cycles of 94°C for 15 s and 55°C for 45 s. A cycle threshold value less than 40 was defined as a positive test. Patient were defined as having a laboratory-confirmed COVID-19 when both targets *(ORF1a/b* and *N* genes) yielded positive results, and repeated tests using another approved RT-qPCR kit were necessary for single-target-positive *(ORF1a/b* or *N* positive) samples.

## RT-LAMP assay

### RT-LAMP Primer Design and Testing

The complete genome sequence of SARS-CoV-2 (GenBank accession number MN908947.3) was aligned and compared the GenBank nucleotide database gene sequences of all species, including other coronaviruses, to identify conserved sequences. A conserved sequence of the *S* gene (nucleotide 22269-22494, No. MN908947.3) was selected as the target to design our RT-LAMP primers because it is highly homologous among various COVID-19 sequences and highly divergent from those of other coronaviruses examined. We designed 4 sets of RT-LAMP primers targeting the SARS-CoV-2 *S* gene sequence (No. MN908947.3) using the online PrimerExplorer V5 software (available at: https://primerexplorerjp/e/). One set of RT-LAMP primers with the best parameters was chosen, including two outer primers, F3 and B3; two inner primers, forward inner primer (FIP) and backward inner primer (BIP); and two loop forward (LF) and backward (LB) primers (Figure S1), all of which synthesized by Invitrogen (Shanghai, China). Primer specificity was verified with a BLAST search of the GenBank nucleotide database via comparisons with other coronaviruses and published SARS-CoV-2 sequences, and the percent mismatch results are presented in Table S2.

*RT-LAMP Assay* For RT-LAMP, each 25-μL reaction comprised 1 μL of 10× primer mix [16 (each) FIP and BIP, 2 μM (each) F3 and B3 primers, 4 μM (each) LF and LB primers]; 2.5 μL of 10× Isothermal Amplification Buffer Pack (New England Biolabs); 4 μL of 10 mM dNTPs; 4 μL of 5 M betaine; 3 μL of MgSO_4_; 2 μL of Bst DNA polymerase (8 U/μL); 1 μL of AMV reverse transcriptase (5 U/μL); 1 μL of 3 mM fluorescent detection reagent (HNB); 5 μL of RNA template and 2.5 μL of 1%o DEPC H_2_O. The reaction mixtures were incubated in a PCR thermocycler or dry bath at 65°C for 35 min. The optimal incubation condition of 65°C for 35 min was determined based on the banding pattern observed after gel electrophoresis and an absorption spectrum analysis of the RT-LAMP reactions (Figure S2). Nontemplate controls (NTCs) were included in each run to ensure the absence of contamination. Positive reactions could be observed by visual color change from purple to blue, fluorescent light in response to UV excitation, or by the laddering pattern of bands after gel electrophoresis.

## Cross-reactivity evaluation of the RT-LAMP assay

Synthesized plasmids of 12 common viral pathogens, SARS, MERS, influenza A H1N1/H3N2, influenza B, human parainfluenza viruses (HPIV-1/2/3), respiratory syncytial virus (RSV-A/B), Epstein-Barr virus, human cytomegalovirus, human mastadenovirus (HAdV-B/E), enterovirus (EB-U/71), human rhinovirus (HRV-2/14/16) and coxsackievirus (CA16), were used to test potential cross-reactivity in the developed RT-LAMP assay. The RT-LAMP products obtained using these plasmid templates were assayed by 3% agarose gel electrophoresis.

## Detection limit of the RT-LAMP assay

To determine the lower detection limit of the RT-LAMP assay, samples from a 10-fold gradient dilution series of synthetic SARS-CoV-2 S gene cDNA (1.5×10^2^ - 1.5×10^-9^ ng/reaction) were used as template in RT-LAMP reactions, and the minimum concentration of the positive reaction was recorded. This dilution series was assayed in parallel by RT-qPCR using primers that targeted the same region of the SARS-CoV-2 genome. The detection limit of the RT-LAMP assay was determined by comparing the lowest concentration of the positive reaction with that obtained by RT-qPCR.

## Multiplex PCR-based next-generation sequencing (NGS)

The samples yielding inconsistent results inconsistent between the RT-LAMP and RT-qPCR assays and those from COVID-19 patients that tested negative by RT-qPCR were further analyzed by multiplex PCR-based enrichment and NGS to detect the SARS-CoV-2 genome. Briefly, total RNA was reverse transcribed to synthesize first-strand cDNA with random hexamers and a Superscript III reverse transcriptase kit (Vazyme, China). Two-step SARS-CoV-2 genome amplification was performed with two pooled mixtures of primer sets (designed by Genskey Medical Technology Co., Ltd.) that were designed to cover the entire SARS-CoV-2 genome. cDNA was mixed with the components of the first PCR following the manufacturer’s instructions. The 2nd PCR was performed using the index primers, and the constructed libraries were sequenced on an Illumina NovaSeq PE 150 platform. Data analysis was primarily performed based on an in-house pipeline produced by Genskey Medical Technology. Raw sequences were quality trimmed and subsequently filtered if shorter than 130 bases using fastp v0.19.5. Sequence reads were first filtered against the human reference genome and then aligned to a reference genome of SARS-CoV-2 (NC_045512.2) using Bowtie v2.2.4. The mapped reads were assembled with SPAdes v3.14.0 with kmers ranging from 19 to 109 to obtain the coronavirus genome sequences.

## Statistical analysis

The sensitivity, specificity, positive and negative predictive values, likelihood ratios and their respective 95% confidence intervals for the RT-LAMP and RT-qPCR assays of nasopharyngeal specimens were calculated, and agreement analysis was computed using kappa concordance coefficients (a value ≥ 0.75 was deemed good) and percentage agreement (≥ 0.9 was considered good). ^15^ Statistical analyses were performed in the R programming environment.

## Ethics statement

Written informed consent was obtained from all participants before the study, and the study was approved by the ethics committee of each participating institution. The analysis was conducted on samples collected during standard COVID-19 tests, with no extra burden on patients.

## RESULTS

### Development of an RT-LAMP assay

As described in the Materials and Methods, in this study we developed a rapid and simple RT-LAMP assay to detect SARS-CoV-2 RNA, where positive reactions resulted in a color change from purple to blue due to decreased magnesium concentration in the presence of extensive Bst DNA polymerase activity, while negative reactions retained the purple color. Figure S3 shows the overall procedure of the RT-LAMP assay. RT-LAMP primers for COVID-19 were specific and had a 9.14-37.56% nucleotide mismatch with SARS, MERS and other coronavirus sequences (Table S2). Furthermore, the a cross-reactivity experiment results demonstrated that the RT-LAMP assay did not cross-react with other human-pathogenic coronaviruses and common viral pathogens, supporting the specificity of this assay for COVID-19 (Figure S4). Dilution experiments with the synthetic SARS-CoV-2 S gene were performed to determine limit of detection (LOD) of RT-LAMP relative to that of the RT-qPCR assay for the detection of SARS-CoV-2 (Figure S5). The observed LOD values for the RT-LAMP and RT-qPCR assays were approximately 1.5×10^-8^ ng per 25-μL reaction solution (i.e., 4.23 copies/reaction) and 1.5×10^-7^ ng/reaction (i.e., 42.3 copies/reaction), respectively. The RT-LAMP assay exhibited a 10-fold higher sensitivity than that of the RT-qPCR assay currently being used in clinical settings, a result that is similar to that of previous LAMP-based assays. ^16–17^

### Characteristics of the subjects

We ultimately collected a prospective cohort of 129 patients from Guangdong Provincial People’s Hospital [Cohort I: 24 COVID-19 patients (37 nasopharyngeal swab samples) and 105 COVID-19 exclusion cases (292 nasopharyngeal swabs)] and an independent cohort of 76 patients from Guangdong Second Provincial General Hospital (Cohort II: 28 COVID-19 with 56 nasopharyngeal swabs and 48 non-COVID-19 patients with 96 nasopharyngeal swabs).

The laboratory-confirmed COVID-19 patients had a median age of 46.5 years (IQR, 31-60 years), 69.23% (36/52) were male, and the majority of the patients reported an exposure history presenting primarily with fever, cough/expectoration, and muscle pain/fatigue (Table 1). Most of the COVID-19 patients (94.23%) were identified as nonsevere cases, and only 3 patients were nonsevere cases on admission. Forty of all 52 COVID-19 patients (76.92%) manifested with chest CT imaging abnormalities, with the most common chest CT patterns being ground-glass opacities (53.85%) and bilateral patchy shadowing (38.46%). The remaining 12 (23.07%) cases showed normal CT images. Twenty-one (40.38%) patients had comorbidities, of whom 15.38% had hypertension and 7.69% had diabetes. Forty (76.92%) patients presented with hematologic abnormalities. The demographic and initial clinical characteristics of the COVID-19 patients in the two respective cohorts are provided in Table 1.

**Table 1.** Clinical characteristics of COVID-19 patients in different cohorts

| Clinical features | Cohort I (n = 24) | Cohort II (n = 28) | All patients (n = 52) |
| --- | --- | --- | --- |
| **Sex (men/ women)** | 16/8 | 20/8 | 36/16 |
| **Age, years, median (IQR)** | 52.50 (30.75-61.00) | 42.50 (31.75-51.50) | 46.50 (31.00-60.00) |
| **Nationality** |  |  |  |
| Chinese | 23 | 27 | 50 |
| African | 1 | 1 | 2 |
| **Exposure history— no. (%)** | 22 (91.67) | 25 (89.29) | 47 (90.38) |
| **Symptoms — no. (%)** |  |  |  |
| any | 23 (95.83) | 25 (89.29) | 48 (92.31) |
| fever | 20 (83.33) | 15 (53.57) | 35 (67.31) |
| cough or expectoration | 16 (66.67) | 6 (21.43) | 22 (59.62) |
| muscle pain or fatigue | 3 (12.50) | 9 (32.14) | 12 (42.31) |
| sore throat | 6 (25.00) | 14 (50.00) | 20 (38.46) |
| shortness of breath | 6 (25.00) | 5 (17.86) | 11 (21.15) |
| diarrhea | 3 (12.50) | 5 (17.86) | 8 (15.38) |
| rhinorrhea | 4 (16.67) | 3 (10.71) | 7 (13.46) |
| headache | 1 (4.17) | 5 (17.86) | 6 (11.54) |
| nausea or vomiting | 1 (4.17) | 3 (10.71) | 4 (7.69) |
| **Radiologic findings— no. (%)** |  |  |  |
| abnormalities on CT | 14 (58.33) | 26 (92.86) | 40 (76.92) |
| ground-glass opacity | 10 (41.67) | 18 (64.29) | 28 (53.85) |
| bilateral patchy shadowing | 12 (50.00) | 8 (28.57) | 20 (38.46) |
| local patchy shadowing | 5 (20.83) | 8 (28.57) | 13 (25.00) |
| interstitial abnormalities | 1 (4.17) | 12 (42.86) | 13 (25.00) |
| **Comorbidities— no. (%)** |  |  |  |
| any | 9 (37.50) | 12 (42.86) | 21 (40.38) |
| hypertension | 4 (16.67) | 4 (14.29) | 8 (15.38) |
| diabetes | 2 (8.33) | 2 (7.14) | 4 (7.69) |
| coronary heart disease | 1 (4.17) | 2 (7.14) | 3 (5.77) |
| bronchitis | 3 (12.50) | - | 3 (5.77) |
| cancer | 2 (8.33) | - | 2 (3.85) |
| hypohepatia | 1 (4.17) | 1 (3.57) | 2 (3.85) |
| chronic obstructive pulmonary disease | - | 1 (3.57) | 1 (1.92) |
| kidney injury | - | 1 (3.57) | 1 (1.92) |
| **Coinfection** |  |  |  |
| any | 9 (37.50) | 3 (10.71) | 12 (23.08) |
| mycoplasma | 9 (37.50) | 1 (3.57) | 10 (19.23) |
| hepatitis B virus infection | - | 1 (3.57) | 1 (1.92) |
| *Mycobacterium tuberculosis* | - | 1 (3.57) | 1 (1.92) |
| influenza A | 1 (4.17) | - | 1 (1.92) |
| **Clinical classification** |  |  |  |
| nonsevere | 22 (91.67) | 27 (96.43) | 49 (94.23) |
| severe | 2 (8.33) | 1 (3.57) | 3 (3.57) |
| **Hematologic abnormalities— no. (%)** |  |  |  |
| any | 20 (83.33) | 20 (71.43) | 40 (76.92) |
| c-reactive protein (>5 mg/L, $\times 10^9$/L) | 14 (58.33) | 12 (42.86) | 26 (50.00) |
| lymphocytes (<1.1 or >3.2, $\times 10^9$/L) | 12 (50.00) | 5 (17.86) | 17 (32.69) |
| hemoglobin (<130 g/L) | 3 (12.50) | 8 (28.57) | 11 (21.15) |
| neutrophils (<3.5 or >9.5, $\times 10^9$/L) | 3 (12.50) | 11 (39.29) | 14 (26.92) |
| leukocytes (<3.5 or >9.5, $\times 10^9$/L) | 4 (16.67) | 5 (17.86) | 9 (17.31) |
| platelets (<125, $\times 10^9$/L) | 4 (16.67) | 3 (10.71) | 7 (13.46) |
| lactate dehydrogenase (>250 U/L) | 5 (20.83) | 2 (7.14) | 7 (13.46) |
| interleukin-6 (>7 pg/mL) | 4 (16.67) | 2 (7.14) | 6 (11.54) |
| **Samples collected for SARS-CoV-2 testing** |  |  |  |
| nasopharyngeal swabs | 37 | 28 | 65 |
| sputum | - | 28 | 28 |

### Diagnostic potential of the RT-LAMP assay for COVID-19 patients and an asymptomatic carrier

We first evaluated the clinical application of the RT-LAMP assay on 329 nasopharyngeal specimens from Cohort I. Of these 329 nasopharyngeal swabs, 35 swabs were confirmed as SARS-CoV-2 positive according to the combined criteria of testing positive by results (28 samples) and NGS confirmation (7 samples) (see Tables 2 and S3 and Figure S6). Thirty-one out of 35 clinically positive samples were determined to be positive using the RT-LAMP assay, and 3 of 294 clinically negative samples were observed to show a positive reaction, which were confirmed to be false-positive reaction by NGS. The performance of the RT-LAMP assay was as follows: sensitivity 88.57% (95% CI 74.05–95.46); specificity 98.98% (97.04–99.65); positive-predictive value 91.18% (77.04-96.95); negative-predictive value 98.64% (96.57-99.47); positive likelihood ratio 86.8 (44.8-168.2); and negative likelihood ratio 0.12 (0.07-0.19) (Table 2). Compared with the RT-qPCR assay, the RT-LAMP assay had a significantly better sensitivity (88.57% vs 80.00%) and comparable specificity (98.98% vs 100%) for the diagnosis of SARS-CoV-2 infection (Table 2). The detection results obtained using the RT-LAMP assay were in good concordance with those obtained using the RT-qPCR assay, with a Cohen’s kappa of 0.89 (0.79-1.00), 100% positive predictive agreement and 98.01% negative predictive agreement. These observations are in line with data reported by in studies by Baek et al. ^16–18^ In addition to exploring the diagnostic potential of RT-LAMP on active COVID-19 patients, we also tested the RT-LAMP assay on an asymptomatic COVID-19 carrier. A 22-year-old female presented to our hospital on January 13 2020, with a 16-year history of congenital heart disease and a one-month aggravation of shortness of breath symptoms. After admission, she tested positive by RT-qPCR for SARS-CoV-2 infection in our hospital without any COVID-19/viral pneumonia clinical symptoms or CT findings. Her oropharyngeal swabs were also sent to the Guangzhou CDC for RT-qPCR double-checking and was confirmed as SARS-CoV-2 infection positive on February 12 and February 15. Respiratory samples were collected throughout her illness from February 11 to March 11 and subjected to parallel RT-LAMP and RT-qPCR assays for SARS-CoV-2 detection (see Figure 2). NGS was simultaneously performed for samples yielding inconsistent results between the RT-LAMP and RT-qPCR assays. The number of positive test results obtained by RT-LAMP was 1.37-fold higher than that observed by RT-PCR (11 vs 8), and 4 RT-LAMP-positive but RT-qPCR-negative samples were verified as SARS-COV-2 positive using NGS (Figure 2 and Table S3). During her hospitalization, the RT-qPCR Ct values fluctuated and became negative after February 26, suggesting that a continuous viral shedding pattern and a decreased viral load over time (data not shown, available upon request).

**Table 2.** Diagnostic performance comparison of RT-LAMP and RT-qPCR for SARS-CoV-2 in different cohorts

| Subjects | Clinically positive samples | Clinically negative samples | Sensitivity % (95% CI) | Specificity % (95% CI) | PPV % (95% CI) | NPV % (95% CI) | PLR % (95% CI) | NLR % (95% CI) | Agreement |
| --- | --- | --- | --- | --- | --- | --- | --- | --- | --- |
| **Cohort I** | **n = 35** | **n = 294** |  |  |  |  |  |  |  |
| RT-LAMP |  |  |  |  |  |  |  |  |  |
| Positive | 31 | 3 | 88.57<br>(74.05-95.46) | 98.98<br>(97.04-99.65) | 91.18<br>(77.04-96.95) | 98.64<br>(96.57-99.47) | 86.80<br>(44.8-168.2) | 0.12<br>(0.07-0.19) | Kappa: 0.89 (0.79-1.00);<br>positive predictive agreement: 100%;<br>negative predictive agreement: 98.01% |
| Negative | 4 | 291 |  |  |  |  |  |  |  |
| RT-qPCR |  |  |  |  |  |  |  |  |  |
| Positive | 28 | 0 | 80.00<br>(64.11-89.96) | 100.00<br>(98.71-100.00) | 100.00<br>(87.94-100.00) | 97.67<br>(95.28-98.87) | - | 0.20<br>(0.15-0.26) |  |
| Negative | 7 | 294 |  |  |  |  |  |  |  |
| **Cohort II** | **n = 46** | **n = 106** |  |  |  |  |  |  |  |
| RT-LAMP |  |  |  |  |  |  |  |  |  |
| Positive | 41 | 1 | 89.13<br>(76.96-95.27) | 99.06<br>(94.85-99.83) | 97.62<br>(87.68-99.58) | 95.45<br>(89.8-98.04) | 94.48<br>(13.23-674.7) | 0.11<br>(0.07-0.16) | Kappa: 0.93 (0.77-1.00);<br>positive predictive agreement: 100%;<br>negative predictive agreement: 96.49% |
| Negative | 5 | 105 |  |  |  |  |  |  |  |
| RT-qPCR |  |  |  |  |  |  |  |  |  |
| Positive | 38 | 0 | 82.61<br>(69.28-90.91) | 100.00<br>(96.50-100.00) | 100.00<br>(90.82-100.00) | 92.98<br>(86.76-96.40) | - | 0.17<br>(0.14-0.22) |  |
| Negative | 8 | 106 |  |  |  |  |  |  |  |

Notes: PPV, positive predictive value; NPV, negative predictive value; PLR, positive likelihood ratio; NLR, negative likelihood ratio.

**Figure 2.**
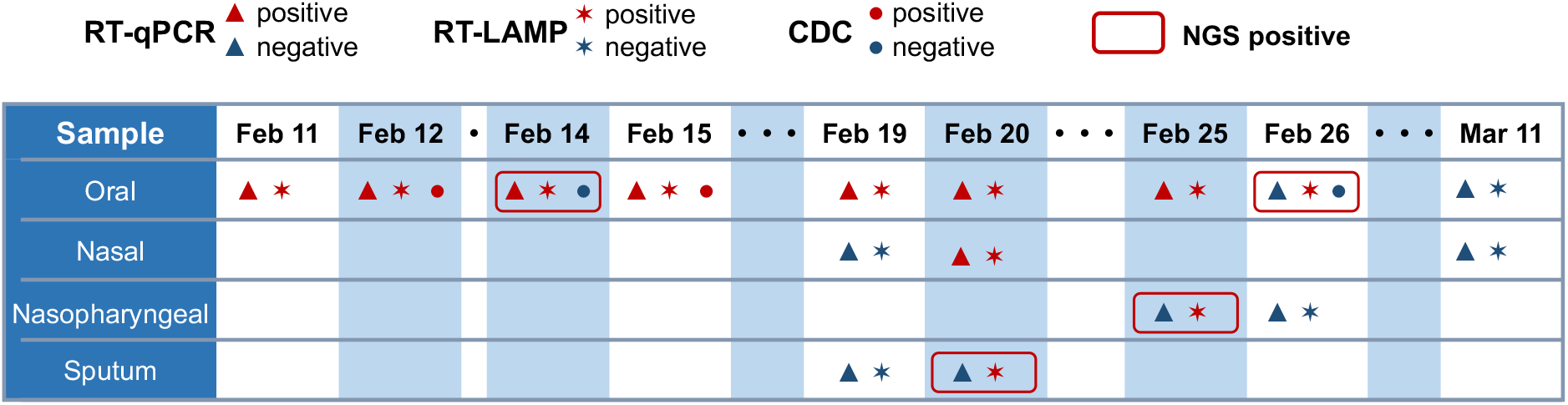
Timeline of detection for an asymptomatic COVID-19 carrier.

This case demonstrated that compared to RT-qPCR, RT-LAMP has a higher sensitivity in detecting SARS-COV-2, particularly for those samples with a low viral load, and also suggested that RT-LAMP can be used for the diagnosis of asymptomatic COVID-19 carriers.

### Validation of RT-LAMP assay

We next validated the RT-LAMP assay on an independent cohort (Cohort II) of 28 COVID-19 patients and 48 COVID-19 exclusion cases. One nasopharyngeal swab and one sputum specimen was collected from every participant in Cohort II. The 152 samples comprised of 46 positive samples (28 swabs and 18 sputum) and 106 negative samples (Tables 2 and S3). Nasopharyngeal swabs from COVID-19 patients showed a higher positive rate than sputum specimens in both the RT-qPCR and RT-LAMP assays [RT-qPCR: swab 71.43% (20/28), sputum 64.29% (18/28); RT-LAMP: swab 78.57% (22/28), sputum 71.43% (20/28)]. The RT-LAMP assay had a sensitivity of 89.13%, whereas that of the RT-qPCR assay was only 82.61%. The specificity of the RT-LAMP assay was roughly equivalent to that of the RT-qPCR assay (99.06% vs 100.00%, Table 2), and the agreement between the two assays was excellent (kappa 0.93 (0.77-1.00), Table 2). These observations corroborate the results obtained from Cohort I as well as previous RT-LAMP findings, ^16–18^ suggesting that the use of RT-LAMP may improve the sensitivity of pathogenic diagnosis for COVID-19.

To further assess whether the RT-LAMP assay was COVID-19 specific, 40 swab specimens from 20 patients with influenza (n = 9) or respiratory viral infections (n = 11, representing *Mycoplasma pneumoniae,* HPIV-1/2/3, RSV-A/B, RSV, and HAdV-B/E) and 20 healthy individuals were assessed using the RT-LAMP assay. No positive results were observed, demonstrating that RT-LAMP-based detection approach can distinguish SARS-CoV-2 with no cross-reactivity for other respiratory viruses, similar to reports in recent studies. ^16–18^

The RT-LAMP assay results reported in this study for SARS-CoV-2 detection in the two cohorts are summarized as follows. The RT-LAMP assay exhibited an overall sensitivity of 88.89% (higher than the 81.48% for RT-qPCR), an overall specificity of 99.00%, high consistency (kappa 0.92) with the RT-qPCR assay, and a median turnaround time less than 1 h from sample preparation to result in the detection of 481 clinical specimens from two cohorts (Figure 3). Additional advantages of RT-LAMP include cost effectiveness, simple operation and visual determination capability, which facilitate SARS-CoV-2 screening in well-equipped labs as well as in the field (Figure 3).

**Figure 3.**
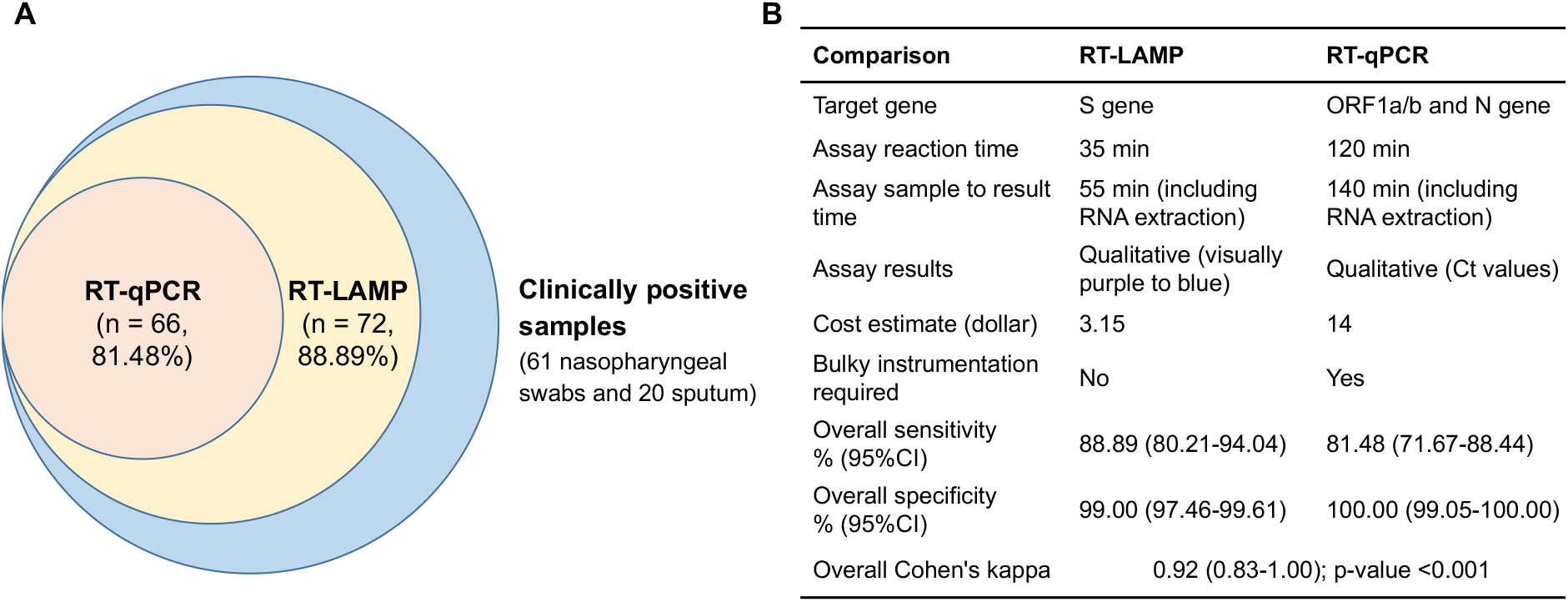
Comparison of the RT-LAMP and RT-qPCR assays for COVID-19 detection in two cohorts. Notes: Eight-one clinical positive samples consists of 35 nasopharyngeal swabs from Cohort I and 46 nasopharyngeal swab and sputum samples from Cohort II, which were determined based on the combined criteria of positive RT-qPCR detection (n = 66) or positive NGS confirmation (n= 15).

## DISCUSSION

Rapid and reliable diagnosis is of particular importance for the containment of COVID-19 outbreaks. In this study, we described a simple and sensitive RT-LAMP approach to rapidly diagnose SARS-CoV-2 infection. The robustness of the present study was demonstrated, as this RT-LAMP assay was useful in the diagnosis of active COVID-19 patients and asymptomatic carriers and was generally not confounded by other respiratory pathogen infections using clinical samples from two hospitals.

Existing methods to detect SARS-CoV-2 are primarily based on RT-qPCR, NGS and IgM and IgG immunological tests. Comparing the results between the RT-LAMP and RT-qPCR assays, RT-LAMP provided a better sensitivity (88.89% vs 81.48%) than RT-qPCR for SARS-CoV-2. This added sensitivity is important in consideration of a significant number of COVID-19 patients that have presented with negative qPCR ^7^ results or the “relapse after negative” phenomenon ^8^ due to potentially large variability between clinical samples, low-virus-titer samples and even disrupted binding of RT-qPCR primers due to variation in the viral genome. ^19^ In this study, we used Bst DNA polymerase that was isolated in-house for the developed RT-LAMP assay, which was demonstrated to have a higher polymerization activity than the commercial Bst DNA polymerase ^20^ and ensured the high sensitivity of this RT-LAMP method. Based on these findings, we propose that the RT-LAMP assay was able to detect viral RNA not only in samples testing positive by RT-qPCR but also in inconclusive samples.

We observed that the RT-LAMP assay was less sensitive and informative than NGS in our study and other literature. ^21–24^ NGS is a robust tool for obtaining extensive genetic information and completing whole-genome sequence, allowing for LOD values as low as 10 copies/mL for SARS-CoV-2 detection and is a reference test for COVID-19, especially for those challenging samples with low viral content. ^2, 23–24^ However, compared to the complex and costly NGS platform, RT-LAMP assays have the advantage of low-threshold of infrastructure, less data processing requirement and cost effectiveness, enabling this user-friendly assay to be immediately deployed in hospitals and communities. The RT-LAMP assay also showed no cross-reactivity with other viruses that manifest similar respiratory diseases such that the specificity of this assay was higher than that reported for IgM- and IgG-based detection methods. ^25^ In addition, we described the accuracy of the RT-LAMP assay in detecting SARS-CoV-2 by determining likelihood ratios. Likelihood ratios are not affected by disease prevalence, and values higher than ten and lower than one strongly support the diagnostic value of a test. ^26^ Based on this metric, the near-patient RT-LAMP assay used in this study is diagnostically useful for COVID-19. Overall, the RT-LAMP assay established in this study could be a powerful complementary method for monitoring massive numbers of exposed individuals as well as aiding with screening efforts in hospitals and public domains, especially in areas with limited laboratory capacities.

In addition, nasopharyngeal swabs from COVID-19 patients had a higher positive rate than sputum specimens in both the RT-qPCR and RT-LAMP assays. Liu et al. ^27^ reported that the detection rate of SARS-CoV-2 RNA in nasopharyngeal swabs was lower than that observed in bronchoalveolar lavage fluid and sputum. This inconsistency is most likely due to poor sputum quality and fluctuations in viral RNA levels during different stages of the disease course. ^28^ Despite this inconsistency, nasopharyngeal swabs are noninvasive and easy to acquire, and evidence has shown that SARS-CoV-2 replicates actively in upper respiratory tissue. ^29^ Therefore, we argue that nasopharyngeal swabs are suitable for the detection of SARS-CoV-2 at an early stage of infection.

We note that four samples from non-COVID-19 cases tested positive in the RT-LAMP assay but negative by RT-qPCR (see Table 1), as reported previously. ^18^ The four false positive results by RT-LAMP were caused by aerosol contaminants, as we retested these samples in another clean room and obtained the expected negative RT-LAMP. Contaminant issues are not rare for nucleic acid testing, even when the best available reference laboratory tests are used. Precautions to prevent cross-contamination or aerosol contaminants during assays are highly recommended, including the use of a spray solution for the elimination of potential RNA fragments and changing gloves frequently. The RNA extraction-free RT-LAMP assay could address this important issue. ^18^ Since this study was completed, the SARS-CoV-2 RT-LAMP test has been optimized further, with the use of lyophilized reagents and the direct detection of SARS-CoV-2 without the need to conduct RNA extraction. This one-step single-tube RT-LAMP assay decreases reaction time and minimizes false positive reactions, making it an ideal POCT for COVID-19 if validated in future studies.

One limitation of our study was the relatively small sample size of positive COVID-19 cases, which has resulted in widened confidence intervals for our estimates of diagnostic accuracy. We tested the samples using RT-LAMP in a blind manner, and the designation of the actual status of SARS-CoV-2 infection in clinical samples was based on a set of combined criteria of RT-qPCR and subsequent NGS confirmation to obviate potential false negative or positive results. We further validated the diagnostic potential of RT-LAMP in another independent cohort with nasopharyngeal swabs and sputum samples. Therefore, despite our small sample size, our study provided sufficient robustness for the RT-LAMP assay.

In summary, in this study, we developed a simple and rapid RT-LAMP assay for SARS-CoV-2 detection and demonstrated its high diagnostic sensitivity and specificity among clinical samples. Our findings suggest that RT-LAMP can be an appropriate auxiliary assay for the diagnosis and epidemiologic surveillance of COVID-19 in different hospital and community settings.

## Data Availability

The supplemental file can be found at the medRxiv website.

